# Response to immune checkpoint inhibition is associated with the gut microbiome in advanced KRAS-mutated non-small cell lung cancer

**DOI:** 10.1101/2023.10.30.23297712

**Authors:** Birgitta I. Hiddinga, Laura A. Bolte, Paul van der Leest, Lucie B.M. Hijmering-Kappelle, Anthonie J. van der Wekken, Ed Schuuring, Ranko Gacesa, Geke A.P. Hospers, Rinse K. Weersma, Johannes R. Björk, T Jeroen N Hiltermann

**Affiliations:** Dept. of Pulmonary Medicine & Tuberculosis, University Medical Center Groningen, University of Groningen, Hanzeplein 1, 9713 GZ Groningen, the Netherlands; Dept. of Gastroenterology & Hepatology, University Medical Center Groningen, University of Groningen, Hanzeplein 1, 9713 GZ Groningen, the Netherlands; Dept. of Genetics, University Medical Center Groningen, University of Groningen, Hanzeplein 1, 9713 GZ Groningen, the Netherlands; Dept. of Pathology, University Medical Center Groningen, University of Groningen, Hanzeplein 1, 9713 GZ Groningen, the Netherlands; Dept. of Medical Oncology, University Medical Centre Groningen, University of Groningen, Hanzeplein 1, 9713 GZ Groningen the Netherlands

## Abstract

**Background:** KRAS-mutated non-small cell lung cancer (NSCLC) is associated with a poor prognosis to standard therapies. Despite advances of immune checkpoint inhibitors (ICIs), not all patients show durable responses. In this study, we aim to identify associations between ICI-response and the gut microbiome in patients with KRAS-mutated NSCLC.

**Methods:** We performed shotgun metagenomic sequencing of stool samples collected before ICI initiation from 33 patients with KRAS-mutated NSCLC. Microbiome composition within (α-diversity) and between samples (β-diversity) was calculated using Shannon diversity index and principal component analysis on Aitchison distances, respectively. A Bayesian logistic-normal regression model (*Pibble*) was implemented to identify associations between gut microbial features and disease control rate (DCR), progression free survival at 12 months (PFS12) and immune related adverse events (irAEs), adjusting for ICI-regimen, metastatic disease stage, age, sex and BMI.

**Results:** Responders were enriched with several saccharolytic species, including *Agathobaculum butyriciproducens, Fusicatenibacter saccharivorans, Bifidobacterium longum* and *Eubacterium ramulus*. Non-responders harbored higher abundances of several Bacteroides and Blautia species. Patients unaffected by irAEs demonstrated higher abundances of biotin and butyrate synthesis pathways. Development of irAEs was associated with higher *Alistipes finegoldii, Bifidobacterium longum* and *Bacteroides uniformis* abundance. No differences were observed between responders and non-responders in Shannon diversity index (*P*=0.69) and overall microbial composition (*P*=0.82).

**Conclusions:** We show gut microbial species and pathways that are differentially abundant between responders and non-responders to ICI in the setting of KRAS-mutated NSCLC. We find overlap with microbial signatures of response to ICI in other tumor types, potentially reflecting tumor-independent microbial mechanisms.

## Introduction

Lung cancer is the leading cause of cancer mortality worldwide. Immune checkpoint inhibition (ICI) has demonstrated significant benefit for patients with advanced non-small cell lung cancer (NSCLC). Recently, pembrolizumab has moved forward as standard of care first line treatment, in NSCLC patients having a PD-L1 tumor proportion score (TPS) > 50% [1]. Nivolumab is standard of care for second and further line treatment of immunotherapy-naïve patients [2]. The Kirsten rat sarcoma viral oncogene homolog (KRAS) mutation is the most frequent genetic alteration found in NSCLC. KRAS mutations are associated with considerable heterogeneity in clinical characteristics and a poor prognosis to standard NSCLC therapies [3]. Immunotherapy seems to be an effective choice in patients with KRAS mutation, in any line of treatment and with better outcomes than chemotherapy [4]. However, responses in most patients are still poor and for up to 80% this treatment will have no favorable effect in terms of long-term survival [5].

The gut microbiome has been recognized as a hallmark of cancer [6]. Mechanisms through which the gut microbiome affects cancer development and progression include eliciting (innate) tumour promoting inflammation as well as escaping (adaptive) immune destruction [6, 7]. Moreover, the gut microbiome has been linked to ICI response, including the development of immune-related adverse events (irAEs), suggesting that characterisation of the gut microbiome may enable a more personalised line of treatment [8, 9, 10]. While most of the evidence comes from melanoma patients, it is not yet clear whether the gut microbiome can serve as a target in patients treated with ICI for different tumor entities such as KRAS-mutated NSCLC [11, 12]. In a French cohort of 338 patients with NSCLC, of which about 40% KRAS-mutated NSCLC, baseline *Akkermansia muciniphila* abundance was associated with increased response rates and overall survival [13].

In this study we investigate the role of the gut microbiome in a cohort of patients treated with anti-PD-1 immunotherapy for advanced KRAS-mutated NSCLC, presenting a specific tumor entity that has not yet been studied in this setting.

## Materials and methods

### Participant selection

From 1^st^ of October 2017 to 1^st^ of December 2019 we enrolled 40 patients with stage IIIB to IVB KRAS-mutated NSCLC (TNM classification of lung cancer, 8th edition) for treatment with ICI as first line treatment (14 patients pembrolizumab, PD-L1 TPS >50%) or after failing first line platinum containing doublet chemotherapy (26 patients nivolumab, independent of PD-L1 score). Patients were treated with nivolumab 3 mg/kg every 2 weeks or pembrolizumab 200 mg flat dose every 3 weeks until progression or intolerable toxicity.

All patients were selected from a cohort of NSCLC patients harbouring a KRAS mutation, as reported previously [14]. Key eligibility criteria are depicted in **Supplementary table 1**. Baseline characteristics, tumour stage and previous treatment are presented in **Table 1**. Antibiotic and proton pump inhibitor (PPI) use within 3 months of commencing ICI were documented.

**Table 1.** Cohort characteristics. Cohort characteristics are presented as mean and standard deviation (SD) for continuous variables and as counts and percentages for categorical variables. Abbreviations: BMI, body mass index; DCR, Disease control rate; ICI, immune checkpoint inhibitor; irAEs, immune-related adverse events; NSCLC, Non-small cell lung cancer; PFS, Progression-free survival; PFS12, Progression-free survival at 12 months; PPI, proton pump inhibitors.

|  | (n=33) |
| --- | --- |
| **Patient characteristics** |  |
| Age (years) at stage IV diagnosis, *mean (SD)* | 64.24 (7.83) |
| Gender, *n (%)* |  |
| female | 18 (55) |
| male | 15 (45) |
| BMI (kg/m<sup>2</sup>), *mean (SD)* | 24.93 (4.53) |
| Performance status, *n (%)* |  |
| PS 0 | 6 (18) |
| PS 1 | 23 (70) |
| PS 2 | 4 (12) |
| Metastatic stage, *n (%)* |  |
| 0 (Stage 3b) | 1 (3) |
| 1a | 10 (30) |
| 1b | 12 (36) |
| 1c | 10 (30) |
| Treated brain metastases, *n (%)* | 3 (9) |
| Liver metastases, *n (%)* | 3 (9) |
| Smoking, *n (%)* |  |
| Current smoker | 4 (12) |
| Former smoker | 29 (88) |
| **Treatment characteristics** |  |
| ICI used, *n (%)* |  |
| Nivolumab (second or further line) | 21 (64) |
| Pembrolizumab (first line) | 12 (36) |
| Antibiotic use at baseline, *n (%)* | 3 (9) |
| PPI use at baseline, *n (%)* | 19 (58) |
| **Outcomes following ICI** |  |
| DCR, *n (%)* | 8 (24) |
| PFS (months), *median (range)* | 1.0 (0-51) |
| PFS12, *n (%)* | 6 (18) |
| irAEs, *n* (%) | 11 (33) |
| Maximum grade irAEs, *n* (%) |  |
| 0 | 22 (67) |
| 1 | 1 (3) |
| 2 | 3 (9) |
| 3 | 4 (12) |
| 4 | 3 (9) |

Of 40 recruited participants two were excluded due to presence of a second primary tumor and not harbouring a KRAS mutation, respectively. Five participants did not collect a faecal sample, leading to 33 patients eligible for analysis.

### Sample collection, DNA extraction and sequencing

Patients received oral and written instructions about the stool sample collection. Stool samples were collected at baseline. The protocol for faecal sample collection and profiling of gut microbiota was previously published [15]. Microbial DNA was isolated with the QIAamp Fast DNA Stool Mini Kit (Qiagen, Germany), according to the manufacturer’s instructions. Metagenomic sequencing was performed at Novogene, China using the Illumina HiSeq 2000 platform. We obtained a total of 7.9 (sd=1.2) Gb with an average of 26.3 (sd=4.0) mil. reads/sample prior to quality control and pre-processing.

### Sample processing

Reads aligning to the human genome (GRCh37/Hg19) were removed using KneadData integrated Bowtie2 tool (V.2.3.4.1), functional profiles were calculated using *HUMAnN3* (V.0.10.0) and the taxonomic composition was evaluated using *MetaPhlAn3*. Microbes and microbial functions that were present in less than 10% of samples and microbes with a relative abundance lower than 0.01% were not included in subsequent analyses. Samples with a sequencing depth below 10□million reads were removed. Arcsine square-root transformations for taxonomic abundances and logarithmic transformation for pathways were used as normalization methods.

### Radiological evaluation and definition of clinical endpoints

Radiological evaluation with CT-scan according to Response Evaluation Criteria in Solid Tumours (RECIST) v1.1 [16] was performed at baseline and every 6 weeks in the first year of ICI treatment, and thereafter every 12 weeks until disease progression.

Clinical endpoints were disease control rate (DCR), progression-free survival at 12 months (PFS12) and development of immune-related adverse events (irAEs).

DCR was defined on the basis of the first two radiological evaluations (at week 6 and week 12) using RECIST v1.1 criteria [16], classifying patients as responders (complete response, partial response, or stable disease) or non-responders (progressive disease). Stable disease was only classified as a response when confirmed at 6 months.

PFS was defined as the time from the first dose of an ICI to the first event; i.e., disease progression or death from any cause, with PFS12 indicating a complete/partial response or disease stability up to at least 12 months following initiation of ICI treatment.

IrAEs during or after ICI treatment were documented using the Common Terminology Criteria for Adverse Events (CTCAE) v5 (**Table 1**) [17]. Side effects of clearly non-immune etiology were excluded.

### Statistical analysis

The statistical tests and terminology are described in **Box 1**. χ2 Tests for categorical variables and Mann-Whitney U test (MWU) for continuous data were performed to calculate differences between responders and non-responders (**Supplementary Table 2**). To test for differences in α-diversity, we computed Shannon diversity index using estimate_richness (…, measures=“Shannon”) from the phyloseq package [18]. To test for differences in β-diversity, we performed a Principal Component Analysis (PCA) on clr-transformed relative abundances using transform(…, transform=“clr”) and ordinate(…, method=“RDA”) from the microbiome and phyloseq package, respectively [18]. To determine which endpoints and variables could significantly explain interindividual variation in the gut microbiome in this cohort, we performed Permutational Multivariate Analysis of Variance (PERMANOVA) on an Aitchison distance matrix produced from species-level clr-transformed relative abundances using the function adonis from the vegan R package (v2.5-7) [19]. The *P* and *R*^*2*^ values were determined by 9999 permutations using all variables in the model.

To identify associations between treatment outcomes and species abundance and metabolic pathways, we implemented a Bayesian logistic-normal linear regression model called *Pibble* from the R package fido [20, 21], which allows for associating covariates to compositional and overdispersed high throughput sequencing data (**Box 1**).

We were particularly interested in the covariates determining whether a likely association existed between the gut microbiome and response to ICI, either DCR (yes/no) or PFS12 (yes/no), ≥ grade 2 irAEs (yes/no), ICI regimen (Nivolumab/Pembrolizumab), and metastatic disease stage (1A, 1B, 1C), also adjusting for age, sex and BMI. Prior to fitting the model, we mean-centered the continuous covariates age and BMI. Furthermore, we used weighted sum/deviation coding (as opposed to treatment coding) which effectively mean-centers categorical covariates, to weight cases and controls by the number of observations [22]. Then, from the fitted model, we calculated the difference in the marginal means for cases vs. controls for each covariate of interest, and then ranked those to determine which microbial features changed the most between cases and controls. We report results at 75% and 90% credible intervals. This means we concluded that a microbial species or pathway is differentially abundant between cases and controls if 75% or 90% of its posterior distribution do not contain zero (i.e. 75% and 90% Bayesian Confidence Level, BCL).

## Results

### Patient characteristics

Clinical and pathological characteristics are summarized in **Table 1**. The median PFS was 1 month (min=0 months, max=51 months, censoring date August 4, 2022), DCR was 24% (8 patients) and PFS12 was 18% (6 patients). Concomitant PPI was 58% and antibiotic use was low (9%). The KRAS G12C mutation was most frequently found (49%). In 11 patients (33%) irAEs occurred, of which 10 were ≥ CTCAE-grade 2. In responders, grade and number of organs affected by irAEs was higher than in non-responders, although not statistically significant (**Supplementary table 2)**.

### Overall gut microbiome composition and diversity

We tested whether the gut microbiome of responders and non-responders exhibited differences in α-diversity and β-diversity and we found no difference between responders and non-responders in the Shannon diversity index neither for microbial species nor pathways in the PFS12 group (**Figure 1**). Similarly, there was no difference between responders and non-responders when using DCR as response measure (both *P=*0.69; **Supplementary figures 1-2**).

**Figure 1:**
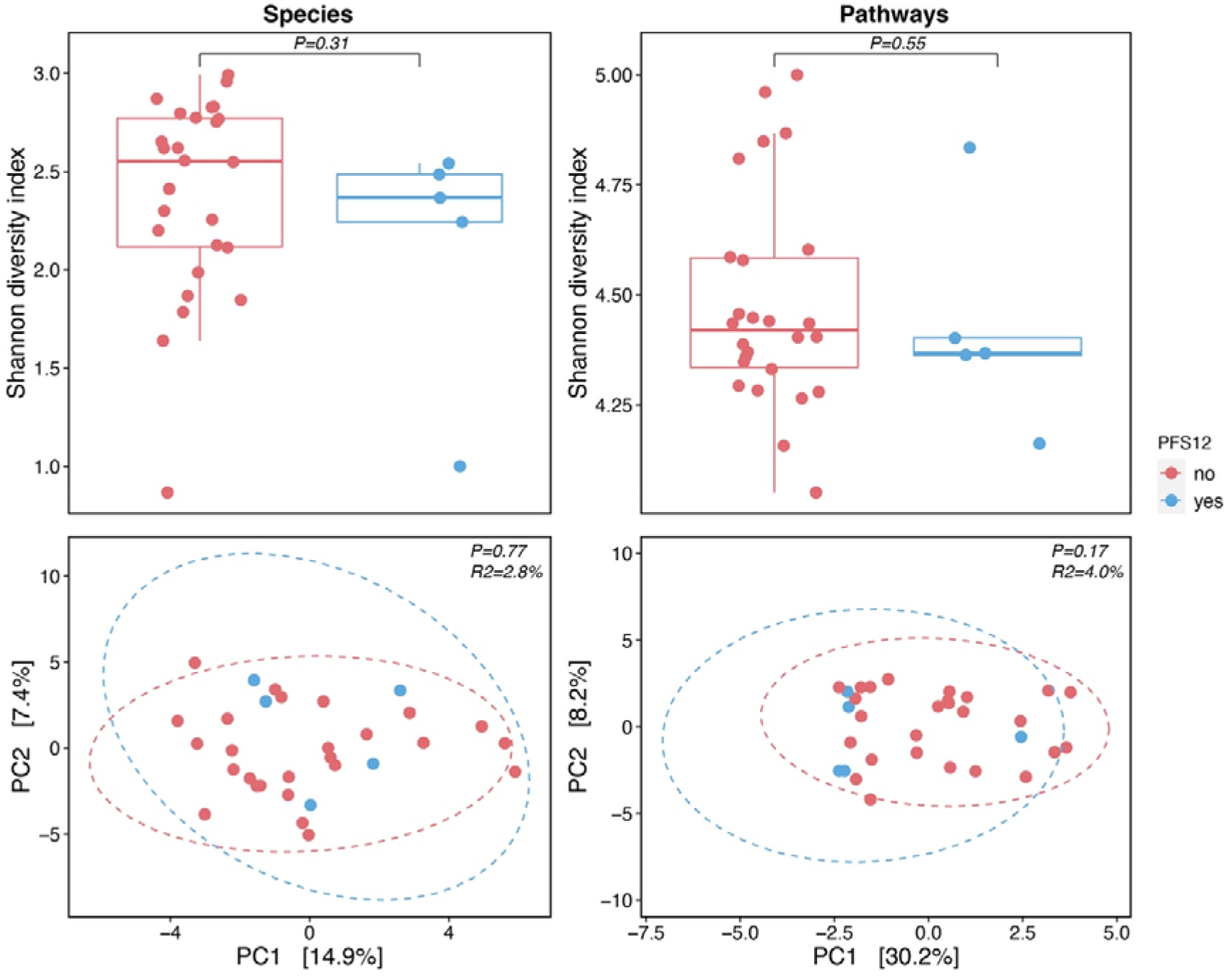
α-and β-diversity between responders and non-responders. Upper panels show α-diversity at the species-(left) and pathway-level (right). α-diversity is computed as the Shannon diversity index (y-axis) for responders (R; blue) and non-responders (NR; red), respectively. Lower panels show species-and pathway-level compositional similarity (β-diversity) between responder and non-responder samples. β-diversity was computed using Aitchison distances. Each eclipse includes 95% of each group’s samples.

We did not find significant differences in microbial species or pathway composition between responders and non-responders for either PFS12 nor DCR (**Supplementary table 3**). Thereafter, we tested whether patients who developed irAEs exhibited differences in α-and β-diversity compared to those who were resistant to irAEs. Similarly to response to ICI, we found no differences between these two patient groups in terms of the Shannon diversity index for species, pathways, nor for microbial species or pathway composition (**Supplementary figures 1-2, Supplementary table 3**). In the PERMANOVAs, we also included and tested whether ICI regimen, metastatic disease stage, sex, age and BMI explained variation in gut microbiome composition. For species composition, we found that metastatic disease stage, and sex were the variables explaining most variation in both the DCR and PFS12 model. For microbial pathway composition we found that response (DCR and PFS12), ICI regimen, metastatic disease stage, and age explained the largest percent variation (between 4 and 6%). However, none of the variables reached statistical significance (**Supplementary table 3**).

### Differential abundance analysis

#### Responders show enrichment of short chain fatty acids (SCFA)-producers

Responders were enriched in several saccharolytic species involved in the synthesis of short chain fatty acids (SCFA), including *Agathobaculum butyriciproducens, Clostridium leptum, Bifidobacterium longum, Eubacterium ramulus* and *Fusicatenibacter saccharivorans* (**Figure 2A)**. Furthermore, responders showed higher relative abundances of *Akkermansia muciniphila* and SCFA-producers *Alistipes putredinis* and *Alistipes finegoldii* compared to non-responders, although these associations showed a wider credible interval (**Figure 2**). At pathway level, responders showed a higher abundance of pathways involved in synthesis of biotin (BIOTIN-BIOSYNTHESIS-PWY) and butyrate (PWY-5676; PWY-5022) (**Figure 2B**).

**Figure 2:**
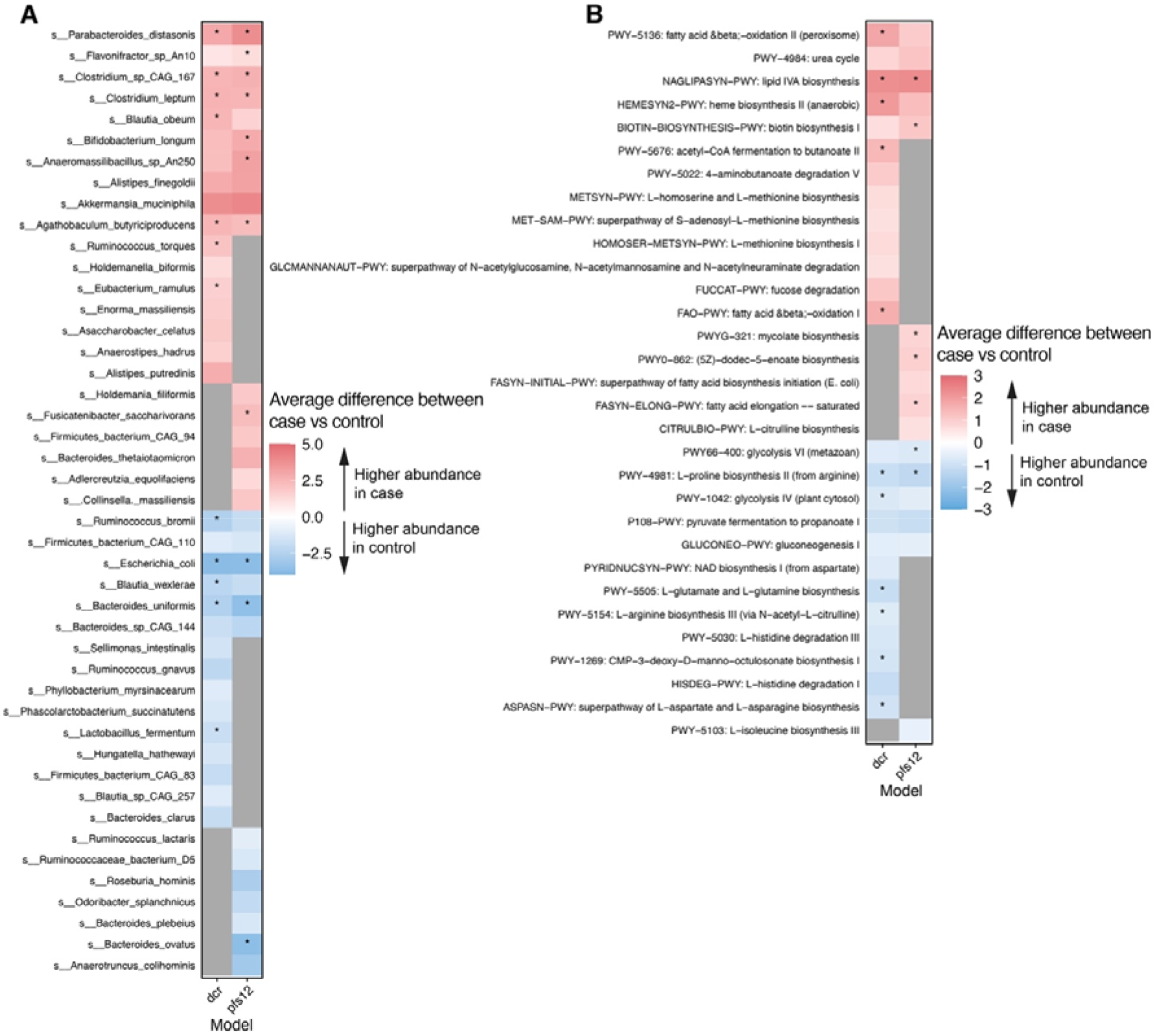
Differential abundance analysis. Differentially abundant microbial species (A) and pathways (B) between responders (R; red) and non-responders (NR; blue) at 75% Bayesian confidence level. Dots indicate microbial features that were differentially abundant at 90% Bayesian confidence level. Color strength indicates the effect size.

#### Responders show higher abundance of immunogenic pathways

In contrast to the aforementioned health-associated microbial features, we also observed that responders had higher abundances of pathways involved in lipopolysaccharide (LPS) and heme synthesis (NAGLIPASYN; PWY-5136; FAO-PWY; HEMESYN2-PWY, **Figure 2B**). These pathways are generally regarded as pro-inflammatory.

#### Higher abundance of Bacteroides species in non-responders

Non-responders were enriched in several species belonging to the Bacteroides (*Bacteroides (B*.*) sp CAF 44; B. clarus; B*.*wexlerae; B. uniformis; B. plebeius; B. ovatus*) and Blautia genus (*Ruminococcus gnavus; Blautia sp. CAG 257*) in both the DCR and the PFS12 model. Non-response was also associated with a higher abundance of *Escherichia coli* and several amino acid synthesis pathways (**Figure 2**).

#### Biotin and SCFA synthesis pathways enriched in patients unaffected by irAEs

Patients who did not develop irAEs showed higher abundances of *Anaerotruncus colihominis* and *Hungatella hathewayi* (that were associated with non-response) and *Anaerostipes hadrus* (**Supplementary figure 3**). These patients further showed an enrichment of pathways involved in the synthesis of biotin and SCFA or precursors of SCFA (BIOTIN-BIOSYNTHESIS-PWY; PWY-5676; PWY-5022) and fatty acid synthesis (FASYN-INITIAL-PWY; FASYN-ELONG-PWY); **Supplementary figure 4**).

On the other hand, patients who developed irAEs exhibited higher abundances of *Alistipes finegoldii* and *Bifidobacterium longum* (that were enriched in responders) and *Bacteroides uniformis* (enriched in non-responders; **Supplementary figure 3**). Development of irAEs was associated with microbial synthesis of several amino acids (lysine, arginine, proline, isoleucine, asparagine; **Supplementary figure 4**) partially overlapping with pathways seen enriched in non-responders (**Figure 2B**).

#### Comparison of different ICI-regimen and metastatic disease stages

Finally, we compared gut microbial abundances in different treatment settings, including metastatic disease stage and treatment line or agent. We observed higher abundances of *Bacteroides sp. CAG 144* and *Escherichia coli* in those treated with first-line pembrolizumab compared to those treated with second- and further line nivolumab (**Supplementary Figure 3)**. Patients with an earlier metastatic disease stage (M1a) harbored higher abundances of *Bifidobacterium* and *Eubacterium spp*., biotin synthesis (BIOTIN-BIOSYNTHESIS-PWY) and starch degradation (PWY-2723) pathways, whereas the higher metastatic disease stage M1c was associated with higher abundances of a pathway involved in LPS synthesis (NAGLIPASYN-PWY; **Supplementary Figures 5-6**).

## Discussion

In this study we profiled the gut microbiome composition and function in a homogeneous cohort of patients treated with anti-PD-1 immunotherapy for advanced KRAS-mutated NSCLC. We identified distinct gut microbial features of response and non-response to treatment, while correcting for important clinical confounders such as ICI-type, metastatic disease stage and the development of irAEs. In line with earlier studies, response-associated microbiome features were not reflected at the whole microbiome level by common β-diversity metrices [23].

### Overlap in microbial signatures of response with other tumour types and geographies

We found that responders are enriched in SCFA producing species compared to non-responders. SCFA producing species have previously been associated with healthy host phenotypes [11, 15, 24, 25]. SCFA-producers such as *Bifidobacteria* [23, 10], *Faecalibacterium prausnitzii* [9, 26] have been consistently associated with ICI-response across several tumour types including melanoma and renal cell carcinoma [8, 27], geographies [23, 28], and treatment regimen [13, 29].

Similarly, *Akkermansia muciniphila* has been repeatedly associated with increased overall survival and response rates to ICI in cohorts from different countries, such as in advanced NSCLC patients from France [8, 13] and Poland [30], and melanoma patients from the Netherlands [31], the UK and Spain [23]. In our cohort we found increased *Akkermansia muciniphila* in responders, but not statistically significant, probably due to the small sample size. We also identified two *Alistipes* species to be enriched in responders, in line with the findings in melanoma and NSCLC patients [8, 26, 32].

At the level of predicted metabolic pathways, we observed an enrichment of biotin synthesis in responders as well as in patients unaffected by irAEs. Microbial pathways for the synthesis of biotin, as well as other B-vitamins, have been reported to be enriched in those protected from ICI-induced colitis [33].

Non-responders showed higher abundances of the species belonging to the Blautia genus and the Bacteroides genus. These species have been previously shown to be associated with chronic diseases such as IBD, diabetes mellitus and cardiovascular diseases and long-term diets rich in animal protein and saturated fat [15, 24, 25]. Our observation aligns with the findings of a recent cross-cancer meta-analysis at which *Bacteroides* were relatively underrepresented in ICI-responders across different tumor types including NSCLC, melanoma, hepatic and renal cell carcinoma [8]. *Bacteroides clarus* in particular, has been consistently associated with non-response to ICI [23].

### Differences to NSCLC cohorts from other geographies

Overall, there has been heterogeneity in the microbial species associated with response across different cohorts [23], owing to regional differences such as diets, including concomitant medication use, and to methodological confounders such as limited sample sizes, metastatic disease stage or ICI-type not considered. In contrast to our findings, a study from China found higher abundances of Bacteroidea such as *Bacteroides massiliensis* in NSCLC patients who showed a partial response after anti-PD-1 therapy [32]. Interestingly, previous studies have shown biphasic effects for the *Bacteroides* genus: while the *Bacteroides* genus has been associated with negative efficacy of anti-PD-1 blockade, in line with our study [9], some *Bacteroides* species (*B. fragilis, B. thetaiotaomicron)* have been shown to increase efficacy when anti-CTLA-4 blockade was used [29]. In another study from China, NSCLC patients treated with nivolumab showed an enrichment of *Alistipes, Bifidobacterium* and *Prevotella*, whereas *Ruminococcaceae* was associated with non-response [34]. Two studies from Japan in ICI-treated NSCLC patients found yet another set of species associated with response, mainly *Ruminococcaceae* and *Agathobacter* [35] and *Lactobacillus, Clostridium* and *Syntrophococcus* [36], whereas non-responders were enriched in *Bilophila, Sutterella* and *Parabacteroides*. Another reason for the observed differences could be that, while the patients in our cohort all harboured a KRAS mutation, previous studies conducted in NSCLC did not look at these patients separately. Larger studies across different geografies are needed to further elucidate the role of the gut microbiome in NSCLC.

### Higher abundance of inflammatory pathways in responders

While previous studies suggest a health-associated gut microbiome profile in responders to ICI [37], including higher abundances of SCFA-producers, we also observed an enrichment of microbial functions that are generally considered “pro-inflammatory” or immunogenic. The precise mechanisms between the gut microbiome and immunotherapy still have to be elucidated [38]. Our findings suggest that different microbial mechanisms are at play that could help promote an anti-cancer immune response during ICI treatment, challenging the concept of predominantly healthy microbiome signatures associated with response. Given the role of SCFA producing species in the fermentation of fiber, our results support a potential benefit of fiber-rich diets and unsaturated fatty acids to improve outcomes of ICI therapy [39, 40].

### Strengths and Limitations

To our knowledge this is the first study associating outcomes to ICI with the gut microbiome composition that is conducted exclusively in NSCLC patients harbouring a KRAS mutation, a tumor entity that has been hard-to-target by standard NSCLC therapies in the past. A limitation of the study lies in its sample size and studying only a single time point (pre-treatment). Future multinational studies across different tumor entities with longitudinal profiling of the gut microbiome are needed to confirm the role of the identified species as potential biomarkers or treatment targets to improve response to ICI.

## Conclusions

In a homogeneous cohort of patients with a KRAS-mutated NSCLC, we identified microbial species and pathways associated with response to ICI, and the development of irAEs. We find overlap in gut microbial species and functions associated with ICI-response in other tumour types such as melanoma, that may reflect tumour-independent microbial mechanisms. Specifically, we identified an enrichment in species involved in the fermentation of fiber and production of SCFA in responders, supporting a potential benefit of fiber-rich diets to synergize with ICI. Non-responders harbored a higher abundance of species belonging to the *Bacteroides* genus. The findings support the notion that the gut microbiome could be an interesting target to improve outcomes in NSCLC patients treated with ICI.

## Supporting information

STROBE-checklist-v4-combined

Suppl_Table_3_PERMANOVA

## Data Availability

All relevant data supporting the key findings of this study are available within the article and the supplementary files. Codes used for generating the microbial profiles are publicly
available at [https://github.com/WeersmaLabIBD/Microbiome/blob/master/Protocol_metagenomic_pipeli
ne.md]. All statistical analysis scripts are written in R and can be found here:
https://github.com/WeersmaLabIBD/Microbiome

https://github.com/WeersmaLabIBD/Microbiome/blob/master/Protocol_metagenomic_pipeline.md

## Acknowledgements

The authors would like to acknowledge the funding of the SEERAVE foundation. We thank ms. B.H. Jansen for logistical and laboratory support. We thank all participants of the study for their contribution.

## Box 1.

**Statistical glossary**

α**-diversity:** Quantifies the number of microbial species within each sample. To test for differences in α-diversity, we computed the Shannon diversity index.

**Shannon diversity:** A measure of α-diversity which penalizes rare species.

β**-diversity:** Quantifies the similarity/dissimilarity between two different samples. To test for differences in β*-*diversity, we performed a Principal Component Analysis (PCA) on clr-transformed relative abundances.

**Aitchison distance:** The euclidean distance between samples calculated on species relative abundances after center log-ratio transformation (clr). This distance is considered the gold standard for high throughput sequencing data.

**Principal Component Analysis (PCA):** A dimensionality reduction technique that is used to reduce the size of a large dataset into a smaller one that keeps most of the information. The result of this analysis is typically visualized in an ordination plot.

**Bayesian logistic-normal regression model:** Statistical models that allow associating compositional and overdispersed high throughput sequencing data (such as microbiome data) with covariates. In the *Pibble* model, regression coefficients are ranked to determine which microbial features changes the most between cases and controls with statistical significance achieved through Bayesian inference. Importantly, the rankings produced from relative abundances are identical to the rankings produced by absolute abundances.

**Permutational multivariate analysis of variance (PERMANOVA)**: Non-parametric multivariate statistical permutation test. Distance-based method to test which variables could significantly explain interindividual variation in the gut microbiome composition. The test statistics directly use the distance matrix to partition β-diversity into different sources of variation.

**Microbial species**: Groups of microorganisms that share common genetic and phenotypic characteristics. A species is a group of similar organisms (strains) within a genus. Microbial species can play important roles in various biochemical pathways and metabolic processes, such as the breakdown of fiber through fermentation, which can produce energy and metabolic byproducts, short-chain fatty acids.

**Microbial pathways**: Refer to specific biochemical processes and metabolic pathways that are carried out by microorganisms. Predicted metabolic functions of gut microbiota are based on their annotated genome and can be captured by whole shutgun metagenomics sequencing.

## Tables and Figures

**Supplementary table 1** Inclusion criteria

**Supplementary table 2** Descriptive statstics

**Supplementary table 3** PERMANOVA analysis (*Excel file*)

**Supplementary figure 1** Alpha and beta diversity at the species-level

**Supplementary figure 2** Alpha and beta diversity at the pathway-level

**Supplementary figure 3** Species-level comparison of irAEs and different ICI-regimen

**Supplementary figure 4** Pathway-level comparison of irAEs and different ICI-regimen

**Supplementary figure 5** Species-level comparison of different metastatic disease stages

**Supplementary figure 6** Pathway-level comparison of different metastatic disease stages

## References

1. Reck M, Rodríguez-Abreu D, Robinson AG, et al. Updated Analysis of KEYNOTE-024: Pembrolizumab Versus Platinum-Based Chemotherapy for Advanced Non-Small-Cell Lung Cancer With PD-L1 Tumor Proportion Score of 50% or Greater. J Clin Oncol. 2019 Mar 1;37(7):537–546. doi: 10.1200/JCO.18.00149. Epub 2019 Jan 8. PMID: 30620668.

2. Borghaei H, Gettinger S, Vokes EE, et al. Five-Year Outcomes From the Randomized, Phase III Trials CheckMate 017 and 057: Nivolumab Versus Docetaxel in Previously Treated Non-Small-Cell Lung Cancer. J Clin Oncol. 2021 Mar 1;39(7):723–733. doi: 10.1200/JCO.20.01605. Epub 2021 Jan 15. PMID: 33449799.

3. Salgia R, Pharaon R, Mambetsariev I, Nam A, Sattler M. The improbable targeted therapy: KRAS as an emerging target in non-small cell lung cancer (NSCLC). Cell Rep Med. 2021 Jan 19;2(1):100186. doi: 10.1016/j.xcrm.2020.100186. PMID: 33521700; PMCID: PMC7817862.

4. Kim JH, Kim HS, Kim BJ. Prognostic value of KRAS mutation in advanced non-small-cell lung cancer treated with immune checkpoint inhibitors: a meta-analysis and review. Oncotarget 2017.

5. Skoulidis F, Goldberg ME, Greenawalt DM, et al. STK11/LKB1 Mutations and PD-1 Inhibitor Resistance in KRAS-Mutant Lung Adenocarcinoma. Cancer Discov. 2018 Jul;8(7):822–835. doi: 10.1158/2159-8290.CD-18-0099. Epub 2018 May 17. PMID: 29773717; PMCID: PMC6030433.

6. Liu C, Zheng S, Jin R, et al. The superior efficacy of anti-PD-1/PD-L1 immunotherapy in KRAS-mutant non-small cell lung cancer that correlates with an inflammatory phenotype and increased immunogenicity. Cancer Lett 2020;470:95–105.

7. Hanahan D. Hallmarks of Cancer: New Dimensions. Cancer Discov. 2022 Jan;12(1):31–46. doi: 10.1158/2159-8290.CD-21-1059.

8. Routy B, Le Chatelier E, Derosa L, et al. Gut microbiome influences efficacy of PD-1-based immunotherapy against epithelial tumors. Science. 2018 Jan 5;359(6371):91–97. doi: 10.1126/science.aan3706. Epub 2017 Nov 2. PMID: 29097494.

9. Gopalakrishnan V, Spencer CN, Nezi L, et al. Gut microbiome modulates response to anti-PD-1 immunotherapy in melanoma patients. Science. 2018 Jan 5;359(6371):97–103. doi: 10.1126/science.aan4236. Epub 2017 Nov 2. PMID: 29097493; PMCID: PMC5827966.

10. Matson V, Fessler J, Bao R, et al. The commensal microbiome is associated with anti-PD-1 efficacy in metastatic melanoma patients. Science. 2018 Jan 5;359(6371):104–108. doi: 10.1126/science.aao3290. PMID: 29302014; PMCID: PMC6707353.

11. Gui Q, Li H, Wang A, Zhao X, Tan Z, Chen L, Xu K, Xiao C. The association between gut butyrate-producing bacteria and non-small-cell lung cancer. J Clin Lab Anal. 2020 Aug;34(8):e23318. doi: 10.1002/jcla.23318. Epub 2020 Mar 29. PMID: 32227387; PMCID: PMC7439349.

12. Nagasaka M, Sexton R, Alhasan R, et al. Gut microbiome and response to checkpoint inhibitors in non-small cell lung cancer - A review. Crit Rev Oncol Hematol. 2020 Jan;145:102841. doi: 10.1016/j.critrevonc.2019.102841.

13. Derosa L, Routy B, Maltez Thomas A, et al. Intestinal Akkermansia muciniphila predicts clinical response to PD-1 blockade in patients with advanced non-small-cell lung cancer. Nat Med 2022;28:315–324. doi: 10.1038/s41591-021-01655-5.

14. van der Leest P, Hiddinga B, Miedema A, et al. Circulating tumor DNA as a biomarker for monitoring early treatment responses of patients with advanced lung adenocarcinoma receiving immune checkpoint inhibitors. Mol Oncol. 2021 Nov;15(11):2910–2922. doi: 10.1002/1878-0261.13090. Epub 2021 Sep 25. Erratum in: Mol Oncol. 2022 Jan;16(1):310. PMID: 34449963; PMCID: PMC8564646.

15. Gacesa R, Kurilshikov A, Vich Vila A, et al. Environmental factors shaping the gut microbiome in a Dutch population. Nature 2022;604:732–739.

16. Eisenhauer EA, Therasse P, Bogaerts J, et al. New response evaluation criteria in solid tumours: revised RECIST guideline (version 1.1). Eur J Cancer. 2009 Jan;45(2):228–47. doi: 10.1016/j.ejca.2008.10.026.

17. Common Terminology Criteria for Adverse Events (CTCAE) version 5.0 https://ctep.cancer.gov/protocoldevelopment/electronic_applications/docs/ctcae_v5_q uick_reference_5x7.pdf Assessed 9Dec 2022.

18. McMurdie PJ, Holmes S. Phyloseq: an R package for reproducible interactive analysis and graphics of microbiome census data. PLoS One. 2013 Apr 22;8(4):e61217. doi: 10.1371/journal.pone.0061217. PMID: 23630581; PMCID: PMC3632530.

19. Oksanen J, Simpson GL, Blanchet FG, Kindt R, Legendre P, Minchin PR, et al. vegan: Community Ecology Package. 2022. Available from: https://cran.r-project.org/web/packages/vegan/index.html.

20. Silverman JD, Roche K, Holmes ZC, et al. Bayesian multinomial logistic normal models through marginally latent matrix-T processes. J Mach Learn Res 2022;23: 1–42.

21. Silverman J, Roche K, Nixon M. fido: Bayesian Multinomial Logistic Normal Regression. 2023. Available from: https://cran.r-project.org/web/packages/fido/index.html.

22. Grotenhuis M, Pelzer B, Eisinga R, et al. When size matters: advantages of weighted effect coding in observational studies. Int J Public Health. 2017;62:163–167. doi: 10.1007/s00038-016-0901-1. Epub 2016 Oct 28. PMID: 27796415; PMCID: PMC5288425.

23. Lee KA, Thomas AM, Bolte LA, Björk JR, de Ruijter LK, Armanini F et al. Cross-cohort gut microbiome associations with immune checkpoint inhibitor response in advanced melanoma. Nat Med. 2022 Mar;28(3):535–544. doi: 10.1038/s41591-022-01695-5.

24. Nogal A, Valdes AM, Menni C. The role of short-chain fatty acids in the interplay between gut microbiota and diet in cardio-metabolic health. Gut Microbes 2021;13:1–24. doi: 10.1080/19490976.2021.1897212. PMID: 33764858; PMCID: PMC8007165.

25. Vich Vila A, Imhann F, Collij V, et al. Gut microbiota composition and functional changes in inflammatory bowel disease and irritable bowel syndrome. Sci Transl Med. 2018;10:eaap8914. doi: 10.1126/scitranslmed.aap8914. PMID: 30567928.

26. Chaput N, Lepage P, Coutzac C, et al. Baseline gut microbiota predicts clinical response and colitis in metastatic melanoma patients treated with ipilimumab. Ann Oncol. 2019 Dec 1;30(12):2012. doi: 10.1093/annonc/mdz224. Erratum for: Ann Oncol. 2017 Jun 1;28(6):1368-1379. PMID: 31408090.

27. Frankel AE, Coughlin LA, Kim J, et al. Metagenomic Shotgun Sequencing and Unbiased Metabolomic Profiling Identify Specific Human Gut Microbiota and Metabolites Associated with Immune Checkpoint Therapy Efficacy in Melanoma Patients. Neoplasia 2017;19:848–855. doi: 10.1016/j.neo.2017.08.004. Epub 2017 Sep 15. PMID: 28923537; PMCID: PMC5602478

28. Nomura M, Nagatomo R, Doi K, et al. Association of Short-Chain Fatty Acids in the Gut Microbiome With Clinical Response to Treatment With Nivolumab or Pembrolizumab in Patients With Solid Cancer Tumors. JAMA Netw Open. 2020 Apr 1;3(4):e202895. doi: 10.1001/jamanetworkopen.2020.2895. PMID: 32297948; PMCID: PMC7163404.

29. Vetizou M, Pitt JM, Daillere R, et al. Anticancer immunotherapy by CTLA-4 blockade relies on gut microbiota. Science 2015;350:1079–84.

30. Grenda A, Iwan E, Chmielewska I, et al. Presence of Akkermansiaceae in gut microbiome and immunotherapy effectiveness in patients with advanced non-small cell lung cancer. AMB Express 2022;12:86.

31. Wind TT, Gacesa R, Vich Vila A, et al. Gut microbial species and metabolic pathways associated with response to treatment with immune checkpoint inhibitors in metastatic melanoma. Melanoma Res. 2020 Jun;30(3):235–246. doi: 10.1097/CMR.0000000000000656. PMID: 31990790.

32. Fang C, Fang W, Xu L, et al. Distinct functional metagenomic markers predict the responsiveness to anti-PD-1 therapy in Chinese non-small cell lung cancer patients. Front Oncol 2022;12:837525.

33. Dubin K, Callahan MK, Ren B, et al. Intestinal microbiome analyses identify melanoma patients at risk for checkpoint blockade-induced colitis. Nat Commun 2016;7:10391.

34. Jin Y, Dong H, Xia L, et al. The diversity of gut microbiome is associated with favorable responses to anti-programmed death 1 immunotherapy in Chinese patients with NSCLC. J Thorac Oncol 2019;14:1378–89.

35. Hakozaki T, Richard C, Elkrief A, et al. The gut microbiome associates with immune checkpoint inhibition outcomes in patients with advanced non-small cell lung cancer. Cancer Immunol Res 2020:8:1243–50.

36. Katayama Y, Yamada T, Shimamoto T, et al. The role of gut microbiome on the efficacy of immune checkpoint inhibitors in Japanese responder patients with advanced non-small cell lung cancer. Transl Lung Cancer Res 2019;8:847–53.

37. Derosa L, Routy B, Desilets A, et al. Microbiota-Centered Interventions: The Next Breakthrough in Immuno-Oncology? Cancer Discov 2021;11:2396–2412. doi: 10.1158/2159-8290.CD-21-0236. Epub 2021 Aug 16. PMID: 34400407.

38. Weersma RK, Zhernakova A, Fu J. Interaction between drugs and the gut microbiome. Gut 2020;69:1510–1519. doi: 10.1136/gutjnl-2019-320204. Epub 2020 May 14. PMID: 32409589; PMCID: PMC7398478.

39. Bolte LA, Lee KA, Björk JR, et al. Association of a Mediterranean Diet With Outcomes for Patients Treated With Immune Checkpoint Blockade for Advanced Melanoma. JAMA Oncol. 2023 May 1;9(5):705–709. doi: 10.1001/jamaoncol.2022.7753. PMID: 36795408; PMCID: PMC9936383.

40. Simpson RC, Shanahan ER, Batten M, et al. Diet-driven microbial ecology underpins associations between cancer immunotherapy outcomes and the gut microbiome. Nat Med. 2022 Nov;28(11):2344–2352. doi: 10.1038/s41591-022-01965-2. Epub 2022 Sep 22. PMID: 36138151.

