## Supplementary material for "Response to immune checkpoint inhibition is associated with the gut microbiome in advanced KRAS-mutated non-small cell lung cancer"

**Supplementary Table 1.** Key eligibility criteria

| **Inclusion criteria** | **Exclusion criteria** |
| --- | --- |
| - NSCLC stage IIIB to IVB - KRAS-mutation - Age >18 years - Life expectancy >3 months - PD-L1 TPS > 50% (if first line ICI) - ECOG performance score 0-1 - At least 1 measurable lesion by RECIST v1.1 - Adequate bone marrow reserve and organ function - Completion of any prior palliative radiotherapy at least 2 weeks prior to ICI start | - Active brain or leptomeningeal metastases - Previous anti-PD(L)-1 immunotherapy - Previous treatment for ALK rearrangement, EGFR or BRAF mutation - Active, known or suspected autoimmune disease, except for T1DM - Hypothyroidism only requiring hormone replacement - Skin disorders including vitiligo, psoriasis, or alopecia, not requiring systemic treatment. |

Abbreviations: Immune checkpoint inhibition, ICI; Eastern Cooperative Oncology Group, ECOG; Response Evaluation Criteria in Solid Tumours, RECIST; Type 1 diabetes mellitus, T1DM.

**Supplementary Table 2.** Descriptive statistics

|  | **Total**  **(n=33)** | **Responders**  **(n=8)** | **Non-responders (n=25)** | ***P-value*** |
| --- | --- | --- | --- | --- |
| **Patient characteristics** | | | |  |
| Age (years) at stage IV diagnosis, *mean (SD)* | 64.24 (7.83) | 63.38 (8.23) | 64.52 (7.86) | 0.899 |
| Gender, *n (%)* |  |  |  | 0.081 |
| female | 18 (55) | 7 (88) | 11 (44) |  |
| male | 15 (45) | 1 (13) | 14 (56) |  |
| BMI (kg/m^2^), *mean (SD)* | 24.93 (4.53) | 24.45 (4.56) | 25.08 (4.61) | 0.867 |
| Performance status, *n (%)* |  |  |  | 1.000 |
| PS 0 | 6 (18) | 3 (38) | 3 (12) |  |
| PS 1 | 23 (70) | 4 (15) | 19 (76) |  |
| PS 2 | 4 (12) | 1 (13) | 3 (12) |  |
| Metastatic stage, *n (%)* |  |  |  | 0.182 |
| 0 (Stage 3b) | 1 (3) | 1 (13) | 0 (0) |  |
| 1a | 10 (30) | 2 (25) | 8 (32) |  |
| 1b | 12 (36) | 4 (50) | 8 (32) |  |
| 1c | 10 (30) | 1 (13) | 9 (36) |  |
| Treated brain metastases, *n (%)* | 3 (9) | 0 (0) | 3 (12) | 0.748 |
| Liver metastases, *n (%)* | 3 (9) | 0 (0) | 3 (12) | 0.748 |
| KRAS mutation, *n (%)* |  |  |  | 0.805 |
| G12A | 3 (9) | 0 (0) | 3 (12) |  |
| G12C | 16 (49) | 4 (50) | 12 (48) |  |
| G12D | 3 (9) | 1 (13) | 2 (8) |  |
| G12S | 1 (3) | 0 (0) | 1 (4) |  |
| G12V | 5 (15) | 2 (25) | 3 (12) |  |
| G13C | 1 (3) | 0 (0) | 1 (4) |  |
| G13D | 2 (6) | 1 (13) | 1 (4) |  |
| Q22K | 2 (6) | 0 (0) | 2 (8) |  |
| Smoking, *n (%)* |  |  |  | 1.000 |
| Current smoker | 4 (12) | 1 (13) | 3 (12) |  |
| Former smoker | 29 (88) | 7 (88) | 22 (88) |  |
| **Treatment characteristics** | | | |  |
| ICI used, *n (%)* |  |  |  | 0.179 |
| Nivolumab (second or further line) | 21 (64) | 3 (38) | 18 (72) |  |
| Pembrolizumab (first line) | 12 (36) | 5 (63) | 7 (28) |  |
| Number of treatment lines prior to study, *n (%)* |  |  |  | 0.068 |
| 0 | 12 (36) | 5 (63) | 7 (28) |  |
| 1 | 18 (55) | 3 (38) | 15 (60) |  |
| 2 | 3 (9) | 0 (0) | 3 (12) |  |
| Antibiotic use at baseline, *n (%)* | 3 (9) | 0 (0) | 3 (12) | 0.748 |
| PPI use at baseline, *n (%*) | 19 (58) | 4 (50) | 15 (60) | 0.931 |
| irAEs, *n (%)* | 11 (33) | 5 (63) | 6 (24) | 0.114 |
| Colitis, *n (%)* | 3 (9) | 1 (13) | 2 (8) | 1.0 |
| Maximum grade irAEs, *n (%)* |  |  |  | **0.042** |
| 0 | 22 (67) | 3 (38) | 19 (76) |  |
| 1 | 1 (3) | 0 (0) | 1 (4) |  |
| 2 | 3 (9) | 2 (25) | 1 (4) |  |
| 3 | 4 (12) | 1 (13) | 3 (12) |  |
| 4 | 3 (9) | 2 (25) | 1 (4) |  |
| Number of organs affected by irAEs, *n (%)* |  |  |  | **0.016** |
| 0 | 22 (67) | 3 (38) | 19 (76) |  |
| 1 | 6 (18) | 1 (13) | 5 (20) |  |
| 2 | 4 (12) | 3 (38) | 1 (4) |  |
| 3 | 1 (3) | 1 (13) | 0 (0) |  |

Baseline characteristics are presented as mean and standard deviation (SD) for continuous variables and as counts and percentages for categorical variables. Statistics are provided for the total cohort as well as the subset of responders and non-responders defined by DCR. χ2 tests for categorical variables and Mann-Whitney U test (MWU) for continuous data were performed to calculate differences between responders and non-responders. P-values written in bold indicate nominally significant differences between responders and non-responders (P < .05). There were no statistically significant differences between responders and non-responders after multiple hypothesis testing correction (FDR > 0.5). Abbreviations: BMI, body mass index; DCR, Disease control rate; FDR, False Discovery Rate; ICI, immune checkpoint inhibitor; irAEs, immune-related adverse events; PFS, Progression-free survival; PFS12, Progression-free survival at 12 months; PPI, proton pump inhibitors.

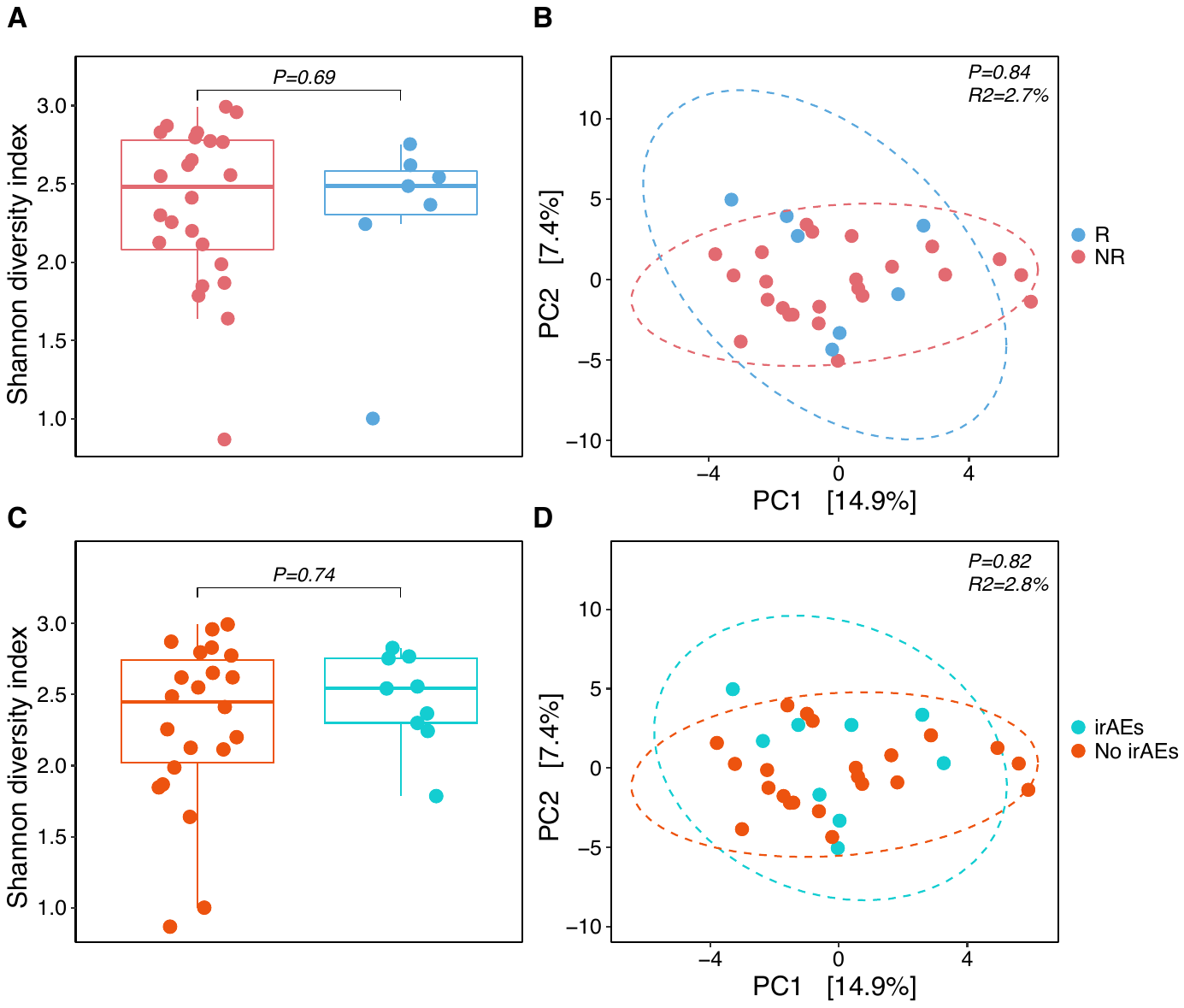

**Supplementary figure 1. Alpha and beta diversity at the species level**

Upper panels show α-diversity (left) and β-diversity (right) for responders (R; blue) and non-responders (NR; red) defined by disease control rate (DCR). α-diversity is computed as the Shannon diversity index (y-axis). Species-level compositional similarity (β-diversity) was computed using Aitchison distances. Each eclipse includes 95% of each group’s samples. Lower panels show α-diversity (left) and β-diversity (right) for patients who developed immune-related adverse events during treatment (irAEs; blue) and those who did no (No irAEs; red).

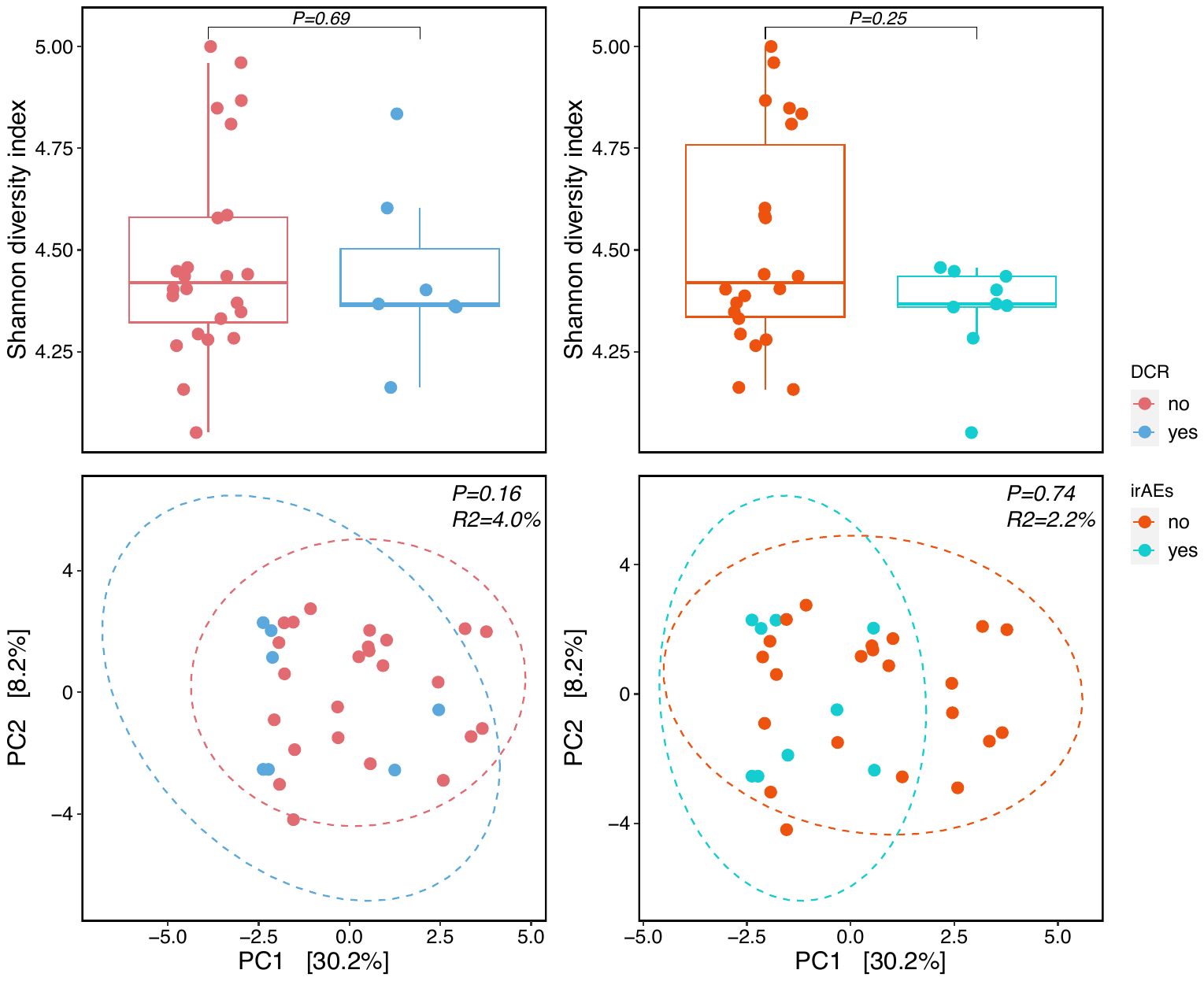

**Supplementary figure 2. Alpha and beta diversity at the pathway-level**

Upper panels show α-diversity (left) and β-diversity (right) for responders (R; blue) and non-responders (NR; red) defined by disease control rate (DCR). α-diversity is computed as the Shannon diversity index (y-axis). Compositional similarity (β-diversity) at the pathway-level was computed using Aitchison distances. Each eclipse includes 95% of each group’s samples. Lower panels show α-diversity (left) and β-diversity (right) for patients who developed immune-related adverse events during treatment (irAEs; blue) and those who did no (No irAEs; red).

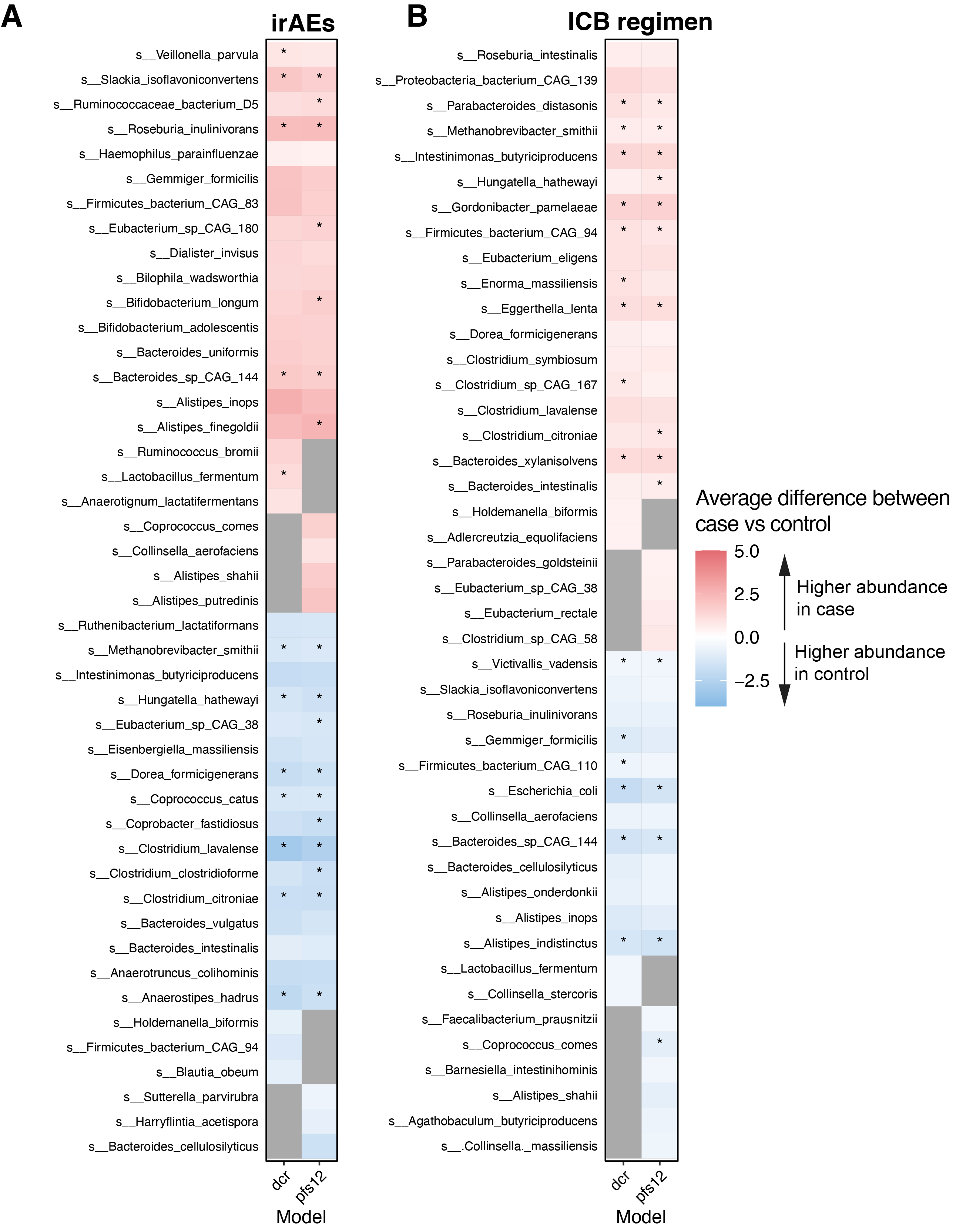
**Supplementary figure 3.** **Species-level comparison of irAEs and different ICI-regimen** Left panel shows differentially abundant microbial species between patients developing irAEs (red) vs. no irAEs (blue) at 75% BCL. Right panel shows differentially abundant species between patients receiving second and further line Nivolumab (red) vs. patients receiving first line Pembrolizumab treatment (blue). Dots indicate microbial features that were differentially abundant at 90% BCL. Color strength indicates the effect size.

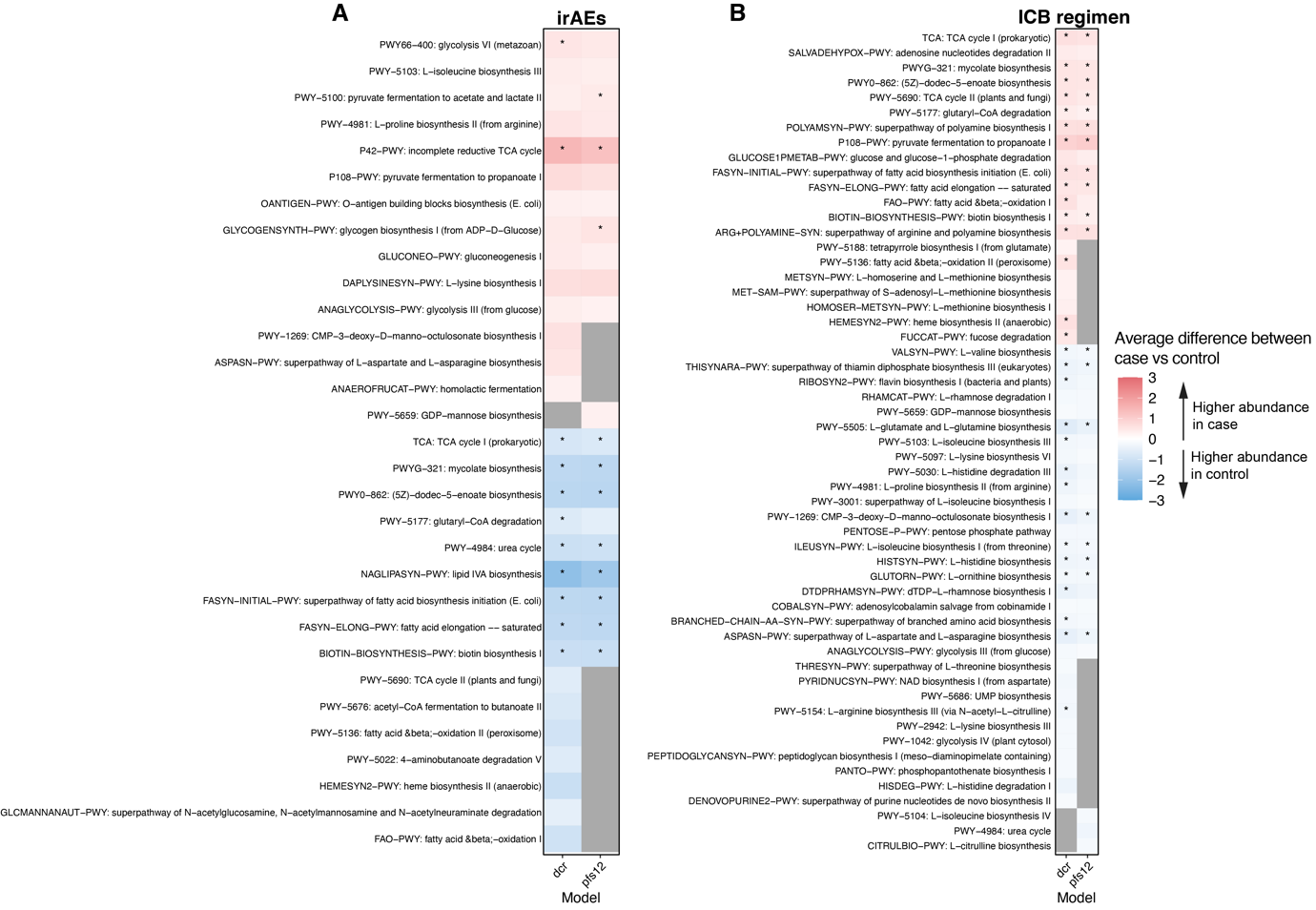

**Supplementary Figure 4.** **Pathway-level comparison of irAEs and different ICI-regimen**

Left panel shows differentially abundant microbial pathways between patients developing irAEs (red) vs. no irAEs (blue) at 75% BCL. Right panel shows differentially abundant pathways between patients receiving second and further line Nivolumab (red) vs. patients receiving first line Pembrolizumab treatment (blue). Dots indicate microbial features that were differentially abundant at 90% BCL. Color strength indicates the effect size.

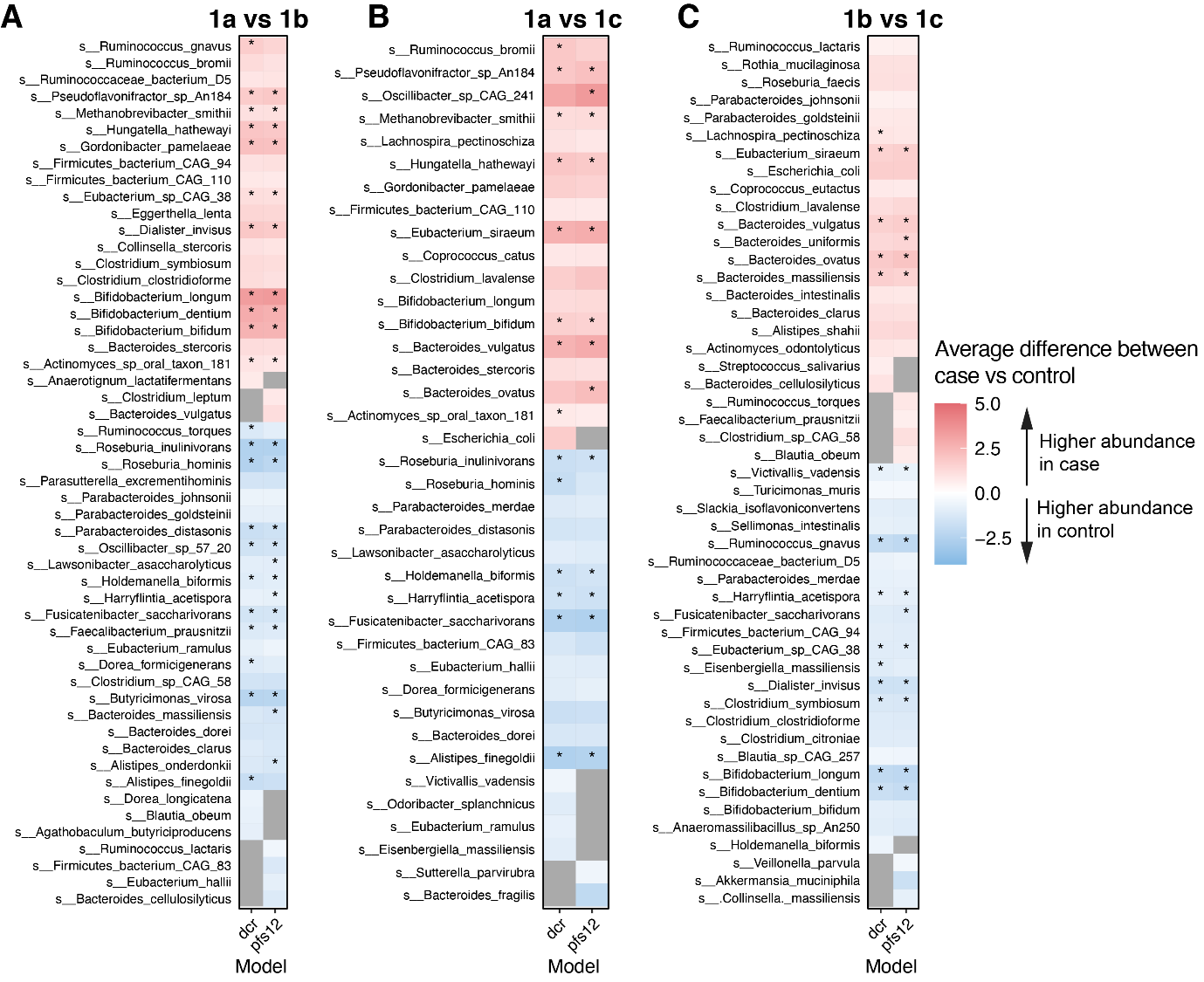

**Supplementary Figure 5. Species-level comparison of different metastatic disease stages**

Differentially abundant microbial species between patients with early disease stages (red) compared to later disease stages (blue) at 75% BCL. Dots indicate microbial features that were differentially abundant at 90% BCL. Color strength indicates the effect size.

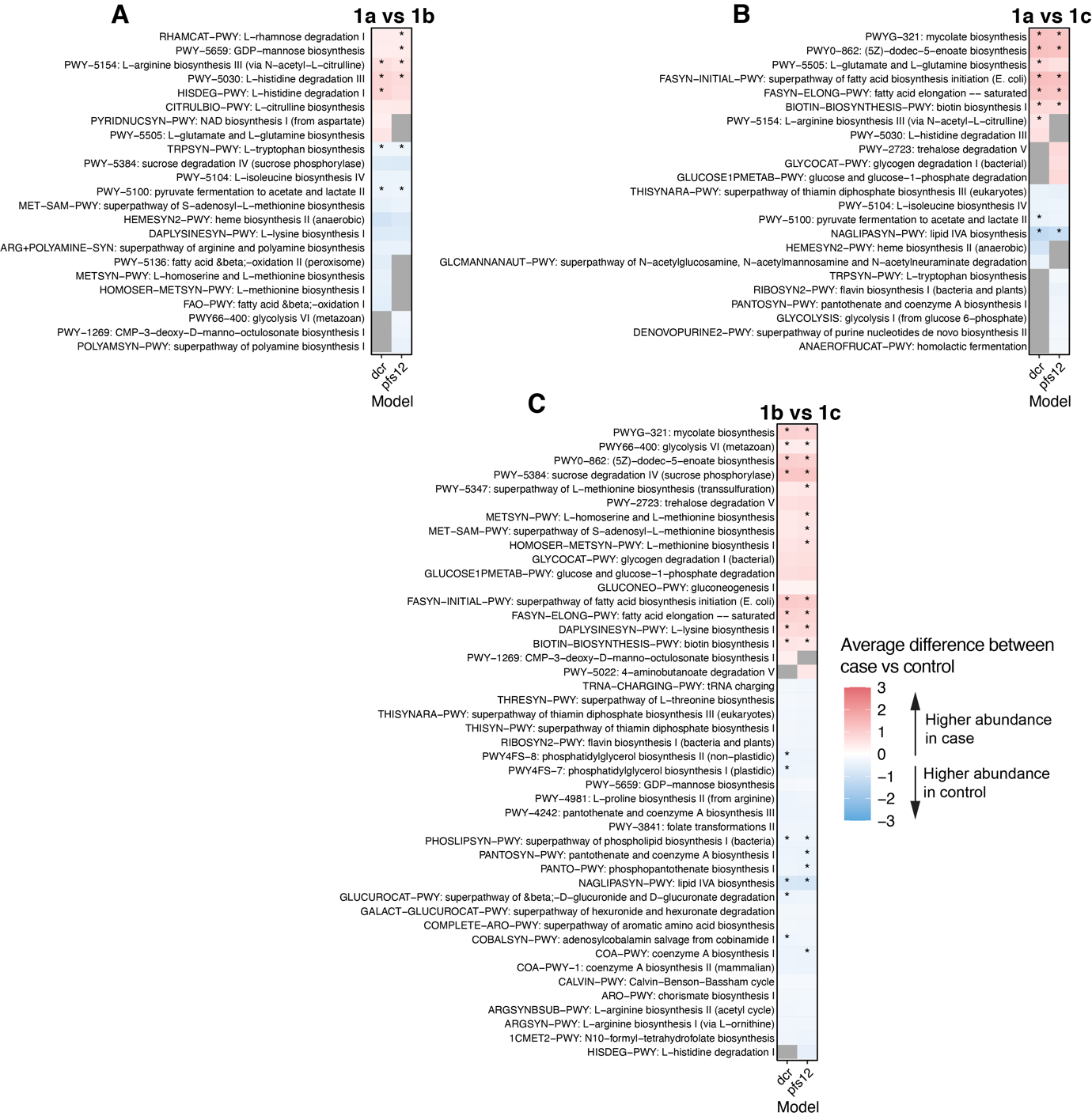
**Supplementary Figure 6**. **Pathway-level comparison of different metastatic disease stages**

Differentially abundant microbial pathways between patients with early disease stages (red) compared to later disease stages (blue) at 75% BCL. Dots indicate microbial features that were differentially abundant at 90% BCL. Color strength indicates the effect size.
